# COVID-19 and children with cancer: Parents’ experiences, anxieties, and support needs

**DOI:** 10.1101/2020.06.11.20128603

**Authors:** Anne-Sophie E Darlington, Jessica E Morgan, Richard Wagland, Samantha Sodergren, David Culliford, Ashley Gamble, Bob Phillips

**Affiliations:** School of Health Sciences, University of Southampton, SO171BJ Southampton, United Kingdom; Centre for Reviews and Dissemination, University of York, YO10 5DD York, United Kingdom; Department of Paediatric Oncology, Leeds Teaching Hospitals NHS Trust, LS1 3EX Leeds, United Kingdom; NIHR Applied Research Collaboration Wessex, University of Southampton, SO171BJ Southampton, United Kingdom; Children’s Cancer and Leukaemia Group, LE1 7GB Leicester, United Kingdom

## Abstract

**Background:** Children with cancer were considered to be extremely clinically vulnerable to severe COVID-19 disease if they were to contract SARS-CoV-2 due to immune suppression as a result of anti-cancer treatment. The aim was to explore experiences, information and support needs, and decision-making of parents with a child with cancer in response to the early phase of the COVID-19 pandemic in the UK.

**Methods:** Parents of a child with cancer completed a survey in April 2020, as the UK moved into a period of ‘lockdown’, with restrictions of movement outside of the home. An online survey was developed to capture parents’ experiences, information and support needs, and decision-making, using closed statements and open text boxes. Descriptive quantitative analyses and qualitative thematic content analysis were undertaken.

**Findings:** 171 parents/caregivers completed the survey. 85% of parents worried about the virus and the majority of parents were vigilant about virus (92%) or cancer symptoms (93.4%). For two-thirds (69.6%) hospital was no longer considered a safe place. Parents worried about their own health (81.1%) and about the child getting the virus from them (89.1%). Eight overarching themes, related to the virus: 1) risk of infection, 2) information, guidance and advice, 3) health care provision, 4) fears and anxieties; or related to lockdown/isolation: 5) psychological and social impact, 6) keeping safe under lockdown, 7) provisions and dependence, and 8) employment and income.

**Interpretation:** This is the first study to report experiences of parents of a child with cancer during the SARS-CoV-2/COVID-19 pandemic. The study demonstrated that the majority of parents are worried about SARS-CoV-2, and worried about transmitting the virus to their child. Hospital was no longer a safe place, and parents were worried about suboptimal cancer care. Parents describe fear and anxiety and the psychological, social and economic impact of isolation as a family.

**RESEARCH INTO CONTEXT**

Evidence before this study
Quarantine due to a pandemic can be traumatic for families. No evidence exists about experiences of families with a clinically vulnerable child with possible increased risk of SARS-CoV-2 infection.

Added value of this study
Insight into high levels of worry about the child with cancer becoming infected by parents, healthcare professionals, or from a hospital visit. Increased vigilance of symptoms, fear and anxiety, psychological impact of isolation, and the need for clear information and guidance.

Implications of all available evidence
Understanding parents’ worries and needs can shape information provision and guidance, and inclusion of vulnerable children in national policy.

## Introduction

Families are worried about SARS-CoV-2 infection; a rapid systematic review of the experience of families under quarantine for recent severe respiratory viruses (SARS-CoV-1, MERS) showed very high levels of traumatic distress.^1^ Parents feel their child with cancer is vulnerable to developing COVID-19.^2^ While cases are few,^3^ and the disease caused by infection has been shown to be relatively mild, surveillance in these groups is encouraged.^4–6^

In the UK, the administrations in England, Wales and Scotland, initially considered children and young people with cancer to be extremely clinically vulnerable to severe COVID-19 disease if they were to contract SARS-CoV-2, due to immune suppression as a result of anti-cancer treatment. They were recommended to ‘shield’ – to remain at home at all times and have no face-to-face contact with anyone outside of their household, except to attend to medical needs. The general population also entered ‘lockdown’ (23 March 2020), with restrictions of movement outside of the home other than for specific designated purposes (i.e. exercise, shopping for essentials, and ‘key workers’ defined as employees who provide vital services maintaining health and essential infrastructure). During this period of time the understanding of the transmissibility of SARS-CoV-2 was uncertain, the nature of the symptoms was evolving, panic buying (stockpiling) was seen and availability and implementation of personal protective equipment (PPE) varied. Through this time, comprehensive and updated advice for parents of children with cancer was compiled and disseminated through national charities and professional organisations in the UK.^2^

Children and young people undergoing treatment face ongoing compromises to the immune system, forcing families to manage infection risks regularly. Decision-making, around continuing treatment, shielding and accessing hospital, under these threatening circumstances needs to be well understood.^7^ This is made even more difficult as new information emerges, for instance with the reporting on multi-system inflammatory disease in children.^8,9^ When information changes, choices change, producing inconsistencies and difficulties.^10–12^ For example, fewer visits by children in emergency departments have already been recorded.^13^

Families of children with cancer have indicated they feel forgotten, with their voice not represented. Existing professional networks of charities, clinicians, academics and parents were mobilised to develop a study to increase our understanding of evolving experiences, information needs and decision-making of these families under these extraordinarily stressful circumstances.

### Aims

To explore experiences, information and support needs, and decision-making of parents of a child with cancer in response to COVID-19.

## Methods

A survey study of parents of a child with cancer, assessing experiences, information and support needs, and decision-making. The findings presented here are part of a larger longitudinal study assessing experiences of parents and children with different paediatric conditions, over time. The survey opened to responses on 6 April 2020 and closed on 4 May 2020, capturing experiences and needs of parents during the first wave, and lockdown, of the COVID-19 pandemic within the UK. The study was approved by the University of Southampton and UK NHS Health Research Authority Research Ethics Committees (IRAS nr. 282176).

### Participants

Parents of a child with cancer aged between 0-18 years able to read and respond in English. Parents were recruited through two principal treatment centres in the UK and through social media, national charities and targeted closed Facebook groups, to minimise the burden on the health system during the pandemic. Electronic consent was obtained before completing the online survey. Approximately 150-200 respondents were intended to be recruited to ensure sufficient numbers of participants to map the range of issues and experiences, identify common issues across them^14–16^ carry out meaningful subgroup analyses, and provide rich data from the open text qualitative data.

### Survey

Survey content was based on currently available literature^17–19^ expert clinician input, and parents. The survey contained the following sections and number of closed statement items: Experiences (n=6), Information (n=7), Decisions (n=7) and Support needs (n=5; Figure 1) Response options for the closed statement items were Not at all, A little, Quite a bit, Very much (except for two conditional questions with Yes/No as response options). Each section started with a free text box for comment before the questions guided the respondent’s thinking. For simplicity, SARS-CoV-2 was referred to as “the virus”. The number of items was purposely small, allowing for rapid analysis and dissemination and increased likelihood of completion. Prior to distributing the survey, feedback from parents was sought about the value and timing of the research, along with detailed questions about the survey in terms of content, phrasing and completeness, and changes were made accordingly.

**Figure 1.**
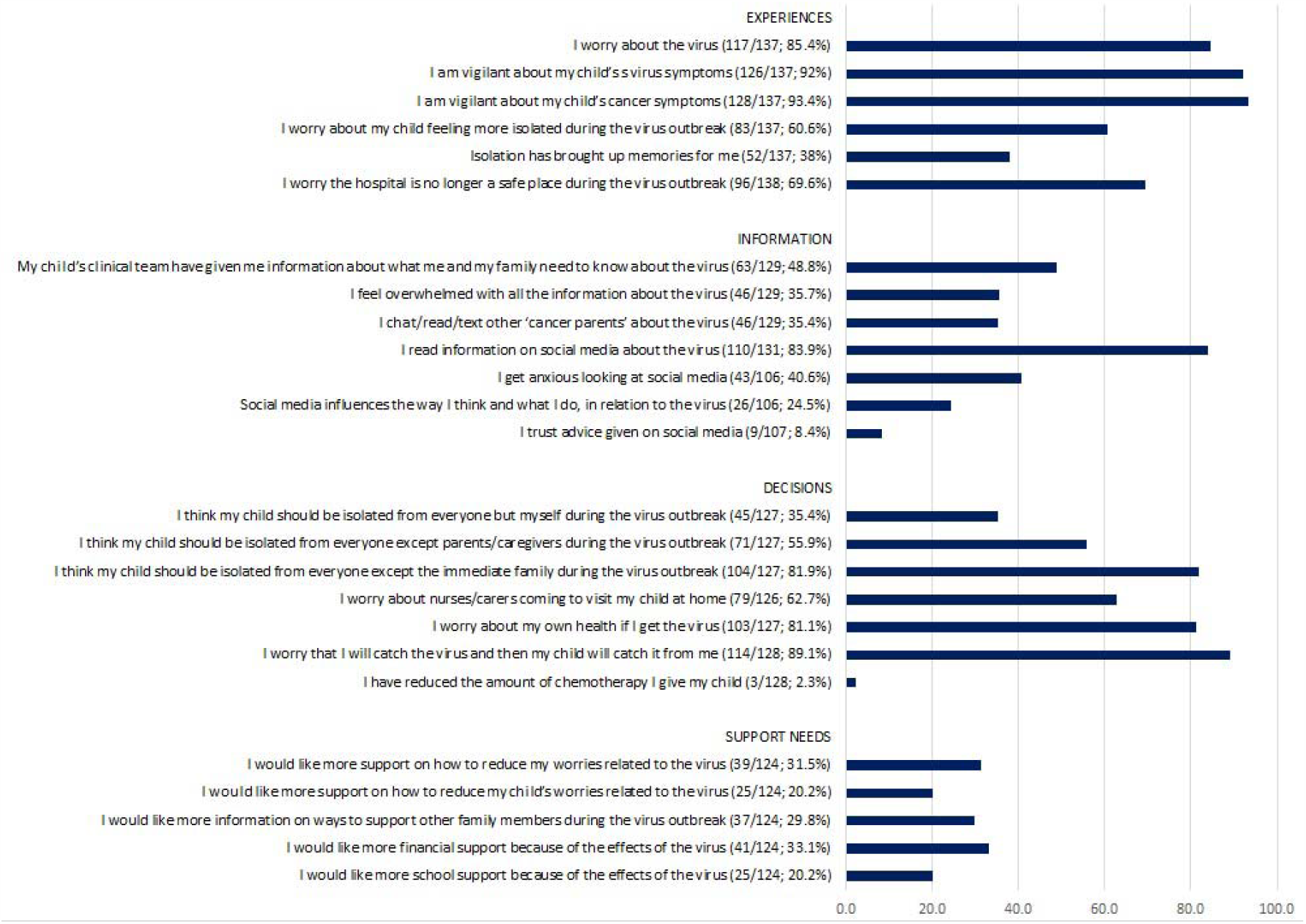
Closed statements percentages (of those who agree Quite a bit or Very much)

### Data analysis

Descriptive statistics were carried out using IBM Statistical Package for Social Science (SPSS)^19^ to summarise the demographic data, and undertake simple descriptive statistics of the closed statement items (collapsing the lowest two response options (Not at all, A little), and the highest two response options (Quite a bit, Very much) into a binary outcome). Subgroup analyses were carried out on an item level, using Chi-squared analyses, according to child’s age (split around the median, age 7 years), treatment (on/off treatment), and diagnosis (Acute Lymphoblastic Leukemia (ALL), solid tumour, Central Nervous System (CNS) tumour, or other). Open text box data were subjected to a thematic content analysis, informed by a three-stage coding process:^20–21^ stage 1) Initial sample of 35 comments were open coded into broad comment categories by two researchers (SS and RW), developing an initial framework, and resolving any conflicts with a third researcher (ASD); stage 2) the framework used to categorise all comments from the data, with further refinement; stage 3) overarching themes were identified from analysis of similarities in the content between categories. Number of comments were counted, to identify weight of themes. Given the overlap in comments to categories the total number of comments did not match the number of participants.

## Results

### Participants

171 respondents completed the survey, of which the majority were mothers (n=143, 83.6%), and nine fathers (Table 1). The child’s median age was 7 years (range 1-24 years). The majority were on treatment (67.3%) and 28.7% were off treatment less than 5 years. The majority of patients were children with ALL (75, 43.9%), and 43 (25.1%) with a solid tumour (Wilms, Rhabdomyosarcoma, Germ Cell tumour, Osteosarcoma, Fibrolamellar Hepatocellular Carcinoma, Neuroblastoma, Retinoblastoma, Ewings Sarcoma, Renal Cell Carcinoma), 12 Lymphoma (7.0%), 11 CNS/brain (6.4%), and six with AML (3.5%).

**Table 1.** Sample characteristics.

| Variables | Values |
| --- | --- |
| Completed by, n (%) |  |
| Mothers | 143 (83.6%) |
| Father | 9 (5.3%) |
| Parent | 9 (5.3%) |
| Other | 4 (2.3%) |
| Missing | 6 (3.5%) |
| Caregiver mean age, years, median (range) | 39 (22-67) |
| Child's age, years, median (range) | 7 (1-24) |
| Child's treatment status, n (%) |  |
| On treatment | 115 (67.3%) |
| Off treatment < 5 years | 49 (28.7%) |
| Off treatment > 5 years | 5 (2.9%) |
| Missing | 2 (1.2%) |
| Diagnosis, n (%) |  |
| Acute Lymphoblastic Leukemia | 75 (43.9%) |
| Solid tumour | 43 (25.1%) |
| Lymphoma | 12 (7.0%) |
| Brain tumour | 11 (6.4%) |
| Acute Myeloblastic Leukaemia | 6 (3.5%) |
| Other | 8 (4.7%) |
| Missing | 16 (9.4%) |

### Closed statement items

A large percentage – those respoding ‘Quite a bit, or Very much-of parents worried about the virus (85.4%), and the majority of parents were vigilant about virus symptoms (92%) or cancer symptoms (93.4%). For two-thirds (69.6%) of the respondents hospital was no longer considered a safe place. Parents received information from their clinical team (48.8%) and accessed information on social media (83.9%), which for some led to feeling anxious (40.6% of those who accessed social media information). Parents isolated their chid from immediate family (81.9%). They worried about their own health (81.1%) and about the child contracting the virus from them (89.1%). The reported worries did not lead to parents stopping or reducing chemotherapy (only 2.3%). The need for support to reduce worries for themselves or others was reported by 20-30% of parents. Group differences in terms of age (0-7 years versus 7-18 years) were found for two items. Parents of younger children were more worried about nurses/carers visiting at home (p=0.001), and more likely to want information on ways to support family members (p=0.002). Parents with a child on treatment were more likely to report that the child should be isolated from everyone except parents/caregivers (p=0.025). No differences were found according to cancer type.

### Open text boxes

#### Experiences open text box

In total, 130 parents (76% of the total) responded to the question about experiences. Overall, the responses to this question covered 38 subthemes (Table 2, including illustrative quotes) which were organised into the following eight overarching themes, related to the virus (four themes) or lockdown and isolation (four themes; Figure 2): <u>Virus</u>: 1) risk of infection, 2) information, guidance and advice, 3) health care provision, and 4) fears and anxieties; <u>Lockdown and isolation</u>: 5)) psychological and social impact, 6) keeping safe under lockdown, 7) provisions and dependence, 8) employment and income (Table 2).

**Table 2.** Themes and subthemes of open text boxes.

| Theme | Subtheme | Number | Quotes |
| --- | --- | --- | --- |
| **VIRUS** |  |  |  |
| **Risk of infection** | **Concern over child's low immunity** | **44** | We are concerned that Covid-19 could get to her easier than the average child |
|  | **Concern over visiting hospitals** | **22** | A place that once was considered safe for our son I now consider to be a great risk due to the risk of catching the virus; The thought of us having to go in overnight is keeping me awake at night. |
|  | Family member has / had Covid-19 | 7 | It's very worrying more so now as my eldest child has symptoms of the virus |
|  | Concern over infection entering the home from parent having to work or shop for provisions | 6 | I am very nervous about going into shops etc. in case I pick something up and take it home. |
|  | Concern over varied approach to wearing PPE | 5 | Some staff are wearing PPE and some are not. |
|  | Vigilance of symptoms | 2 | Extremely on edge about a temperature spike |
| Information, guidance and advice | Limited information/ mixed messaging | 17 | All the information seemed geared at adults not families with vulnerable children. There was a lot of mixed information at the start of the isolation period and far too many grey areas. |
|  | Need for targeted advice and support | 8 | Children within 2 years of transplant are high risk, those people with spleen issues are high risk. Where does this leave our 8 year old? Surely, she cannot be the same low risk as a child who has not had leukaemia, pneumonia, lung fungal infection, possible spleen issues. But we are left on our own in terms of guidance. |
|  | Information regarding child's vulnerability status not issued | 4 | We didn't receive a letter saying "X" was at high risk but when I spoke to somebody at Macmillan they said she was high risk and should be shielding |
|  | Good information from staff | 3 | Luckily our key workers and Leeds children's hospital have given more specific advice appropriate for children with working parents. |
|  | Feel need to seek info from other sources | 2 | Constantly researching the internet looking for case studies for reassurance |
| Healthcare provision | Concern over strained hospital facilities, sub-optimal treatment and care and relapses might be missed | 14 | His next scan is likely to be cancelled so this is causing concern - he has a high-grade tumour which could return quickly so we are worried we could miss a recurrence |
|  | More support required | 6 | Would have preferred some more reassurance and advice from the primary care centre/oncologist as we mainly relied on watching government press conferences. |
|  | New ways of working in the hospital | 7 | Previously, for his chemo appointments, there's a dedicated entrance but now the hospital makes everyone enter through the main entrance which gives an increased possibility in coming into possible coronavirus patients. I understand infection control and some things have to be done but telling us that both parents can be present for 1st chemo session, then that night being informed that wasn't the case and I would have to come alone was quite difficult. |
|  | Priority of Covid-19 over cancer care | 2 | Feel the virus takes priority over everything and we have been left without the same support we had prior to the virus. |
| Fears and anxieties | General expressions of fear | 6 | I think for me it is genuinely the unknown |
|  | Concern over ability to look after child if parent ill or dies | 5 | I am asthmatic and also fear that if I were to get it I couldn't care for her. |
|  | Concern child or parent will die | 4 | What if me and my husband get it and one of us dies, that can't happen |
|  | **Things could be worse** | **3** | On the flip side, we are relishing this time together as a family and so grateful that we are not in the middle of treatment and needing to go to the hospital. |
|  | **Separation if child becomes ill from rest of family** | **3** | I worry that if he catches it I'll have to be in hospital with him away from my other children |
|  | **Child has had/possibly had Covid-19** | **2** | My child has had the virus and it was very mild symptoms. I was very worried about him catching it and thought it would have a bad effect on him but it was very mild |
| **LOCKDOWN AND ISOLATION** |  |  |  |
| **Psychological and social impact** | **Psychological impact on child and family, missing out on life, boredom** | **14** | The isolation has been quite triggering for him, he is bringing up emotions and questions from when he was on treatment. An additional worry, another thing to keep life from being normal. It has restricted any chance of normality during last months of my son's life. We know we only have months, have accepted that, but now we are unable to do the basic things, like go out for coffee, visit grandparents, simple things that bring him pleasure. |
|  | **Parental coping (struggles, strategies used)** | **13** | Panic of not being in control again. As a single parent, it is tough. I need more support from family and friends, that I normally have, but cannot. Feels incredibly lonely. |
|  | **Delayed resumption of normality after treatment** | **9** | We were already isolated from August 19 but we were starting to look forward to that relaxing a bit in the next few months. That has made my daughter really sad. |
|  | **Parallel with cancer treatment isolation** | **7** | As a family we are coping well as isolation is not unusual due to cancer treatment. |
|  | **Missing family and friends** | **5** | It's been hard not seeing family and friends though-this is what has pulled us through our difficult journey and my son is too young to understand why he can only see his grandparents through a window. |
|  | **Impractical nature of social distancing** | **4** | We could not adhere to the ridiculous guidelines set out in the letter of keeping a 2m distance from our young children... caused a lot of stress for families on top of our usual daily stresses. |
|  | **Social and educational development** | **3** | Worry of his social skills being reduced and the long-term impact on him. |
|  | **Missing emotional support for parents from friends and family** | **3** | The virus has taken away my comfort blanket if I feel anxious, I don't have that physical access to family and friends that we did at diagnosis. |
|  | **Use of technology to keep in touch** | **3** | We are using technology to keep in touch with friends and family as that is the hardest bit. |
|  | **Separation from partners/parents/children** | **4** | My husband's work will also not furlough him due to him being a key worker so he has had to move out into the garage for the 12 weeks. |
| **Keeping safe under lockdown** | **Concern over societal compliance in social distancing in society and delayed lockdown** | **10** | I was very worried starting in January that nothing was being done to protect our children especially as it is a brand new disease and nobody knows enough about it. |
|  | **Being on lockdown keeps child safe** | **6** | In some ways because society have restrictions it makes it easier. |
|  | **Concern once restrictions are lifted/ adjustment concerns** | **3** | I worry about re-interacting with society. |
| **Provisions and dependence** | **Difficulty securing provisions (food, cleaning, medication)** | **13** | The hardest with everything is probably the food shopping. We have bought our food online since diagnosis a year ago as we wanted to avoid the shops and now we can't get any delivery slots |
|  | **Lack of priority status** | **4** | One of us is having to go shopping as we are not able to access online shopping as the vulnerable status is in the child's name. |
|  | **Reliance on friends and family to pick up provisions** | **4** | Having to rely on others for everything is quite patronising. |
| **Employment and income** |  | **10** | Worry about finances as both my husband and I have decided to stay home to shield her. |

**Figure 2.**
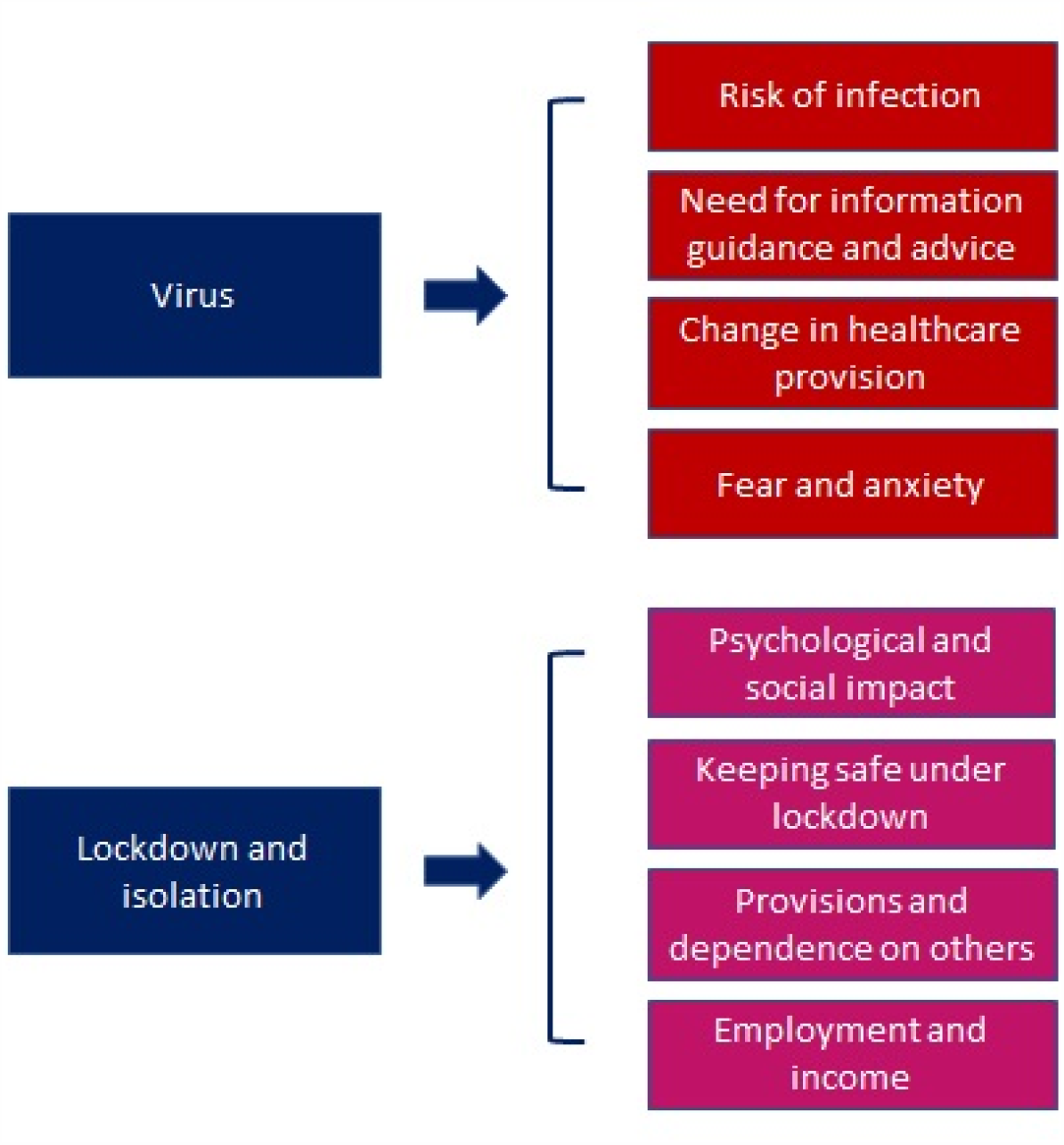
Open text boxes overarching themes.

#### Virus

The largest number of comments (n=44) related to the perceived compromised immune system of their children and their greater susceptibility to the virus. The second largest number of comments (n=22) described safety concerns relating to hospital visits either for outpatient appointments or overnight stays.

Safety of the home environment was felt to be compromised by the virus being brought in by family members who were engaged in a designated ‘essential occupation’ (known as keyworkers in the UK), visiting (community) health professionals or visits to the hospital.

Concerns relating to limited or unclear information from both the hospital and the government were voiced by 17) parents.

Another strong theme (n=14) involved a concern amongst parents that the response to the COVID-19 situation would lead to suboptimal cancer care or had already led to postponed or cancelled clinic appointments, and several parents were concerned that relapses would be missed.

Parents described feeling ‘scared’, ‘terrified’ or ‘petrified’ of the risk of their child becoming infected.

#### Lockdown and isolation

Parents described the psychological impact (n=14), for both themselves and their children, of the emergence of the virus and subsequent lockdown and classification of the child as ‘shielded’, with children struggling mentally, and missing out on life, particularly for those with a limited life expectancy. For parents with children who had recently completed treatment, the lockdown brought with it several frustrations relating to delayed resumption of normality. Parents also described difficulties in coping with the uncertainty of the situation, lack of control, and limited support mechanisms in place which was particularly true for lone parents. Some parents described strategies for coping with the stressful situation, such as the avoidance of or restricted access to news broadcasts and social media.

While under lockdown, parents (n=10) were also concerned about the lack of respect for social distancing shown by some members of the public, the delayed response by the government, and some (n=3) expressed concern about how things might change when restrictions are relaxed.

Access to food home delivery, for families with a child classified as ‘shielded’, and lack of recognition of parents as needing priority status (rather than just the child) (n=13) were concerns.

Financial and employment concerns were also expressed (n=10): parents described having to give up paid work to ‘shield’ their child. In addition, parents expressed frustration in terms of not being eligible for furlough (the government’s Coronavirus Job Retention Scheme, which allowed employers to continue paying wages via a government subsidy).

### Additional free text responses

#### Information

18 parents wanted more information, specifically (n=4) in relation to safety of hospitals, information tailored to children with cancer rather than to adults, information relevant to children with rare cancers and those off treatment, and their child’s level of risk of catching the virus given their particular situation (on or off-treatment) or diagnosis.

For the majority of parents (n=89) charities provided them with information regarding the virus, followed by clinical staff (n=50) and news outlets (n=41).

#### Support

99 parents described a need for additional support related to the provision of more information, specifically more guidance or support from the hospital. The need for information and reassurance surrounding the safety of the hospital environment including the availability and use of personal protective equipment (PPE) as well as testing of staff was further reinforced by 22 parents. In addition, four parents proposed remote contact with hospital staff and services delivered within the community as an option to protect their child.

#### Decisions

Parents were more likely (n=44) to turn to clinical staff when making decisions about their child’s care, while 35 parents described relying on their own judgements based on their knowledge of their child and past experience and 6 were also led by their child when making decisions.

#### Positive

Some parents (n=5) highlighted the positives of the lockdown in terms of bringing the family together and the social restrictions making them ‘feel safe at home’, providing them with a ‘protective bubble’. Some parents (n=3) suggested that things could be worse, or drew comparisons between the isolation imposed during cancer treatment and that of the virus and that they were better equipped than most to face the challenges. In addition, 12 parents took the opportunity to use the survey to communicate their gratitude to the hospital and charities for the care and support they had received.

## Discussion

This is the first study to report experiences of parents of a child with cancer during the SARS-CoV-2/COVID-19 pandemic in the UK. The study has found that the majority of parents were worried about SARS-CoV-2, worried about their own health if they are infected, and worried about transmitting the virus to their child. They described vigilance about SARS-CoV-2 symptoms and cancer-related symptoms, and for a lot of parents the hospital was no longer a safe place during the first month of the lockdown. The qualitative findings show that the threat of SARS-CoV-2 leads to concerns about getting infected and therefore to fear of the hospital and healthcare teams visiting the family at home. Parents got information from their medical team, and almost all parents looked at social media for information, which for some led to feeling anxious. Parents wanted clear information and guidance, which included the shielding policy in the UK. Changes in healthcare provision led some parents to think their care will be suboptimal, and that care for COVID-19 patients was prioritised over that of cancer patients, and that the health service was strained. Parents were anxious about the unknown, about who will look after their child if they get ill (or die). Remarkably, only around one quarter of parents expressed a wish for additional support, and some described how their experience with cancer treatment had made them better prepared for ‘lockdown’ than those without this background.

The lockdown and imposed isolation by the UK government has led to psychological impact in terms of children missing out, feeling bored, missing family and friends, and a delay in social and emotional development, mirroring evidence from studies focusing on young people’s mental health during the pandemic.^23–25^ Parents miss their support network, even though technology is available to connect with others. The designation of children as requiring ‘shielding’ provided particular challenges; the English Government advice described attempting to maintain complete isolation from all other people, even those living in the same household. This lack of age-related nuance may have increased the level of anxiety for some families, particularly around food shopping. Parents struggled with going out to grocery stores, as they did not want to expose themselves to the risk of getting the virus (and thus increasing the chances of the child becoming infected). In common with many people during the lockdown, parents are worried about employment and money; the study group of parents have the additional concern of transmitting the virus to their child.

Uncertainty and lack of clarity in communication were strong themes in the parents’ responses. A clearer, more open, and reasoned account of the various measures being planned and implemented may have assisted with reducing this distress. Much of the uncertainty arose from the true lack of knowledge about the effect of SARS-CoV-2, but few participants reflected this. The all-age signalling of government guidance failed to help families and placed them in an invidious position – to apparently defy the government guideline and risk adversely affecting their child through infection, or follow the guidelines strictly and adversely affect their child with restriction on activity and contact with one parent or siblings.^26^

Reduced attendances in paediatric emergency care facilities with any form of injury or illness were reported.^13^ This, combined with data emerging from this survey on concern about the safety of hospitals, led the charities involved in the research along with local health providers and national paediatric bodies to promote the message of hospitals being ‘safe to attend’. Worries about the possible reduction in anti-cancer therapy were addressed with information co-produced by parents and medical professionals and disseminated through the same routes, explaining the process of contingency planning and the routes to these planned recommendations. The most marked change in care provision was the move to more remote/virtual follow-up appointments, and delay or omission of planned surveillance imaging for patients off treatment. These elements have rarely been shown to have significant survival advantage but contain great emotional weight.^27^

Limitations of the study relate to the bias in the sample - although strenuous efforts were made to widely circulate the survey across children’s cancer interested social media, the respondents may not be representative of the whole population. The responses were mostly from mothers (86%), and the largest group of patients had ALL (46%). While this is disproportionate compared to the diagnoses made in the UK (where it accounts for around one quarter of malignancies), it is also treated for 2-3 years, in comparison with the shorter time frame (under 9 months) of most treatment trajectories. The high proportion of mothers responding is in keeping with surveys about children, as well as the observation mothers being the primary caregiver for the vast majority of children. In addition, parents responding to the survey could have self-selected to represent those parents who were most concerned. Finally, subgroup analyses findings may be based on chance given the number of analyses carried out (n=23×3) and the number of group differences (n=3)

We believe this study demonstrates how the views and experiences of a classically ‘vulnerable’ population can be captured by using existing research networks, and agile governance response and inclusion of patient partners from the commencement of the study. We have found high levels of concern about the consequences of SARS-CoV-2 infection in children with cancer and the consequences of presumed preventative interventions to the children and their families. We propose true uncertainty, coarse recommendations, and a lack of clarity behind decision-making process in national administrations may have worsened these experiences. As the pandemic continues, survey studies such as this will be important in understanding the ongoing experience of families and tuning support and information to their changing needs.

## Data Availability

Anonymised, non-identifiable data will be available for collaborative projects

## Acknowledgements

We would like to thank all parents who contributed their time and experiences to this study.

## References

1. Brooks SK, Webster RK, Smith LE, Woodland L, Wessely S, Greenberg N, Rubin GJ. The psychological impact of quarantine and how to reduce it: rapid review of the evidence. Lancet 2020;395:912–920

2. https://www.cclg.org.uk/Coronavirus-advice

3. Pathak EB, Salemi JL, Sobers N, Menard J, Hambleton IR. COVID-19 in Children in the United States: Intensive Care Admissions, Estimated Total Infected, and Projected Numbers of Severe Pediatric Cases in 2020. Journal of Public Health Management and Practice 2020;26:325–333

4. Kotecha RS. Challenges posed by COVID-19 to children with cancer. Lancet 2020;5:e235

5. Minotti C, Tirelli, Barbieri E3, Giaquinto C, Donà D. How is immunosuppressive status affecting children and adults in SARS-CoV-2 infection? A systematic review. The Journal of Infection 2020 Apr 23.

6. Hrusak O, Kalina T, Wolf J, Balduzzi A, et al. Flash survey on severe acute respiratory syndrome coronavirus-2 infections in paediatric patients on anticancer treatment. European Journal of Cancer 2020;132:11–16.

7. Sung L, Feldman BM, Schwamborn G, Paczesny D, Cochrane A, Greenberg ML, Maloney AM, Hendershot EI, Naqvi A, Barrera M, Llewellyn-Thomas HA. Inpatient versus outpatient management of low-risk pediatric febrile neutropenia: measuring parents’ and healthcare professionals’ preferences. Journal of Clinical Oncology 2004;22:3922–9.

8. PICS society: https://picsociety.uk/wp-content/uploads/2020/04/PICS-statement-re-novel-KD-C19-presentation-v2-27042020.pdf

9. Whittaker E, Bamford A, Kenny J, et al; for the PIMS-TS Study Group and EUCLIDS and PERFORM Consortia. Clinical Characteristics of 58 Children With a Pediatric Inflammatory Multisystem Syndrome Temporally Associated With SARS-CoV-2. JAMA 2020. doi:10.1001/jama.2020.10369

10. Reyna VF, Nelson WL, Han PK, Pignone MP. Decision making and cancer. American Psychologist 2015;70:105–118.

11. Rimer BK, Briss PA, Zeller PK, Chan ECY, Woolf SH. Informed decision making: What is its role in cancer screening? Cancer 2004;101(5 Suppl):1214–1228.

12. Peters E, Klein W, Kaufman A, Meilleur L. Dixon A. More Is Not Always Better: Intuitions About Effective Public Policy Can Lead to Unintended Consequences. Social Issues and Policy Review 2013;7:114–148

13. Isba R,2, Edge R, Jenner R, Broughton E, Francis N, Butler J. Where have all the children gone? Decreases in paediatric emergency department attendances at the start of the COVID-19 pandemic of 2020. Archives of Disease in Childhood 2020 May 6.

14. Hill R. What sample is ‘enough’ in internet survey. Interpersonal computer and Technology Journal 2016;6

15. Gay LR, Diehl PL. Research Methods for Business and Management. 1992 New York: Macmillan.

16. Safdar, N. et al. Research Methods in Healthcare Epidemiology: Survey and Qualitative Research. Infection Control and Hospital Epidemiology 2016;37;1272–1277.

17. Robertson EG, Wakefield CE, Shaw J, et al. Decision-making in childhood cancer: parents’ and adolescents’ views and perceptions. Supportive Care in Cancer 2019;27:4331–4340

18. Morgan JE, Cleminson J, Stewart LA, Phillips RS, Atkin K. Meta-ethnography of experiences of early discharge, with a focus on paediatric febrile neutropenia. Supportive Care in Cancer 2018;26:1039–1050.

19. Morgan JE, Phillips B, Stewart LA, Atkin K. Quest for certainty regarding early discharge in paediatric low-risk febrile neutropenia: a multicentre qualitative focus group discussion study involving patients, parents and healthcare professionals in the UK. BMJ Open 2018;8:e020324.

20. IBM Corp. Released 2019. IBM SPSS Statistics for Windows, Version 26.0. Armonk, NY: IBM Corp.

21. Mason J. Qualitative Researching. 2002, SAGE Publications.

22. Wagland R, Bracher M, Drosdowsky A, et al. Differences in experiences of care between patients diagnosed with metastatic cancer of known and unknown primaries: mixed-method findings from the 2013 cancer patient experience survey in England. BMJ Open 2017;7:e017881.

23. Fegert JM, Vitiello B, Plener PL, Clemens V. Challenges and Burden of the Coronavirus 2019 (COVID-19) Pandemic for Child and Adolescent Mental Health: A Narrative Review to Highlight Clinical and Research Needs in the Acute Phase and the Long Return to Normality. Adolescent Psychiatry and Mental Health 2020;20:14–20.

24. Saurabh K, Ranjan S. Compliance and Psychological Impact of Quarantine in Children and Adolescents due to Covid-19 Pandemic. Indian Journal Pediatrics 2020;29:1–5.

25. Clerici CA, Massimino M, Ferrari A. On the clinical psychologist’s role in the time of COVID-19, with particular reference to experience gained in pediatric oncology. Psychooncology 2020 Jun 5. doi: 10.1002/pon.5418.

26. https://www.itv.com/news/2020-05-06/four-year-old-with-cancer-reunited-with-father-after-seven-weeks-kept-apart-in-lockdown/

27. Morgan JE, Walker R, Harden M, Phillips RS. A Systematic Review of Evidence for and Against Routine Surveillance Imaging After Completing Treatment for Childhood Extracranial Solid Tumors. Cancer Medicine 2020 May 19. doi: 10.1002/cam4.3110.

